# Clinical academic research in the time of Corona: a simulation study in England and a call for action

**DOI:** 10.1101/2020.04.14.20065417

**Authors:** Amitava Banerjee, Michail Katsoulis, Alvina G. Lai, Laura Pasea, Thomas A. Treibel, Charlotte Manisty, Spiros Denaxas, Giovanni Quarta, Harry Hemingway, Joao Cavalcante, Mahdad Noursadeghi, James C Moon

**Affiliations:** Nightingale Hospital, Barts NHS Trust, London, UK; Health Data Research UK, University College London, 222 Euston Road, London, UK; Institute of Health Informatics, University College London, London, UK; Institute of Cardiovascular Science, University College London, London, UK; Department of Cardiology, Ospedale Papa Giovanni XXIII, Bergamo, Italy; Minneapolis Heart Institute, Minneapolis, Minnesota. USA; Division of Infection and Immunity, University College London, London, UK

## Abstract

**Background:** Coronavirus (COVID-19) poses health system challenges in every country. As with any public health emergency, a major component of the global response is timely, effective science. However, particular factors specific to COVID-19 must be overcome to ensure that research efforts are optimised. We aimed to model the impact of COVID-19 on the clinical academic response in the UK, and to provide recommendations for COVID-related research.

**Methods:** We constructed a simple stochastic model to determine clinical academic capacity in the UK in four policy approaches to COVID-19 with differing population infection rates: “Italy model” (6%), “mitigation” (10%), “relaxed mitigation” (40%) and “do-nothing” (80%) scenarios. The ability to conduct research in the COVID-19 climate is affected by the following key factors: (i) infection growth rate and population infection rate (from UK COVID-19 statistics and WHO); (ii) strain on the healthcare system (from published model); and (iii) availability of clinical academic staff with appropriate skillsets affected by frontline clinical activity and sickness (from UK statistics).

**Findings:** In “Italy model”, “mitigation”, “relaxed mitigation” and “do-nothing” scenarios, from 5 March 2020 the duration (days) and peak infection rates (%) are 95(2.4%), 115(2.5%), 240(5.3%) and 240(16.7%) respectively. Near complete attrition of academia (87% reduction, <400 clinical academics) occurs 35 days after pandemic start for 11, 34, 62, 76 days respectively – with no clinical academics at all for 37 days in the “do-nothing” scenario. Restoration of normal academic workforce (80% of normal capacity) takes 11,12, 30 and 26 weeks respectively.

**Interpretation:** Pandemic COVID-19 crushes the science needed at system level. National policies mitigate, but the academic community needs to adapt. We highlight six key strategies: radical prioritisation (eg 3-4 research ideas per institution), deep resourcing, non-standard leadership (repurposing of key non-frontline teams), rationalisation (profoundly simple approaches), careful site selection (eg protected sites with large academic backup) and complete suspension of academic competition with collaborative approaches.

## Introduction

The pandemic SARS-CoV-2 virus (causing the disease, COVID-19) is unprecedented in its impact on individuals, populations and health systems(1). Since the first cases in Wuhan, China in November 2019, every country has been affected(2-3) but with wide variations in the ability and capacity to respond, with only half estimated to have operational readiness(4). Countries hit later can benefit and learn from acquired knowledge and experience of preceding countries. As part of this response, the research effort is crucial for development, testing and adoption of effective preventative and treatment(5,6).

A Pubmed search (November 2019 to April 2020), using terms “coronavirus” and “COVID-19” showed 2206 and 1604 articles respectively, suggesting swift global research mobilisation. However, the publication mix shows the vast majority are reviews, opinions and commentary rather than formal research. Many publications on COVID-19 are not clinically led, and many are not directly clinically informed. “Learning is difficult in the midst of an emergency”(7), but our ability to deliver timely, high-impact clinical research, relevant to patients and populations, is critical across the academic spectrum(8), from “bench to bedside to big data”, whether basic biology, repurposed and novel therapeutic approaches, vaccines or modelling. Obstacles to and strategies for delivering research during a pandemic are poorly characterised.

Anecdotally, many countries have a baseline shortage of clinical academics in translational science(9) and many leading pathfinder health institutions are within major international transport hubs (London, Madrid, New York), which are affected early in the pandemic. Lockdowns close university departments and funding bodies, with alternative funding sources (charities, philanthropy) hit by stockmarket falls and competing demand. Frontline remoteness impedes communication of urgency to decision makers, themselves usually selected for process delivery rather than dynamic adaptability. Critical researchers with relevant virology/immunological/intensive care knowledge are drawn in to local or national clinical responses. Other academic staff most likely to redeploy to COVID-19 research self-select for immediate response roles(10) with universities prioritising repurposing to frontline care(11). High disease rates, required self-isolation periods(12,13) and distractions of remote working degrade the focus needed to create new or repurposed research delivery structures.

We therefore wanted to understand the pandemic research process and describe early lessons. Our aims were to: (i) model potential impact of the pandemic on clinical academic capacity in England relating to COVID-19; and (ii) develop evidence-based recommendations to inform the optimal scientific response to COVID-19.

## Methods

### Cases and excess deaths related to COVID-19 in the UK

Based on our previous analysis of COVID-19 cases and excess deaths in England(14), we considered four scenarios of government interventions associated with different levels of population infection rates: 80% (“*do-nothing*”), 40% (“*relaxed mitigation*”), 10% (“*mitigation”*) and 6% (“*Italy model*”), since “partial suppression”(1%) and “full suppression”(0.001%) were no longer feasible. The analyses of excess deaths used data in a cohort design with prospective recording and follow-up from the Clinical reseArch using LInked Bespoke studies and Electronic health Records (CALIBER) open research platform with validated, reusable definitions of several hundred underlying conditions(15, 16), linking electronic health records(EHR) from different data sources (via UK unique individual identification data, NHS numbers): primary care (Clinical Practice Research Datalink-GOLD), hospital care (Hospital Episodes Statistics), and death registry (Office of National Statistics). Approval was via the Independent Scientific Advisory Committee (16_022R) of the Medicines and Healthcare products Regulatory Agency in the UK in accordance with the Declaration of Helsinki. Key variables were population infection rate, background mortality risk based on underlying conditions, and relative risk (RR) of mortality associated with COVID-19. We used real-time data until 7 April 2020 for the number of confirmed cases and deaths(17).

### Simulation study for population infection rate and infection growth rate

We designed and implemented a simple stochastic model to predict number of new cases in the population. Since the number of new cases are proportional to the active cases of the previous date (see Web appendix), we used official data from 10 April and calculated the ratio 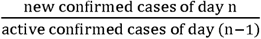 from 5 March onwards. We explored four different scenarios of growth of the infection curve, reflecting different government policies (do-nothing, relaxed mitigation, mitigation and the Italy model), from April 10 (day 36 in our study which coincides with the date of the analysis) until day 250, see Web Table 1.

**Web table 1.**
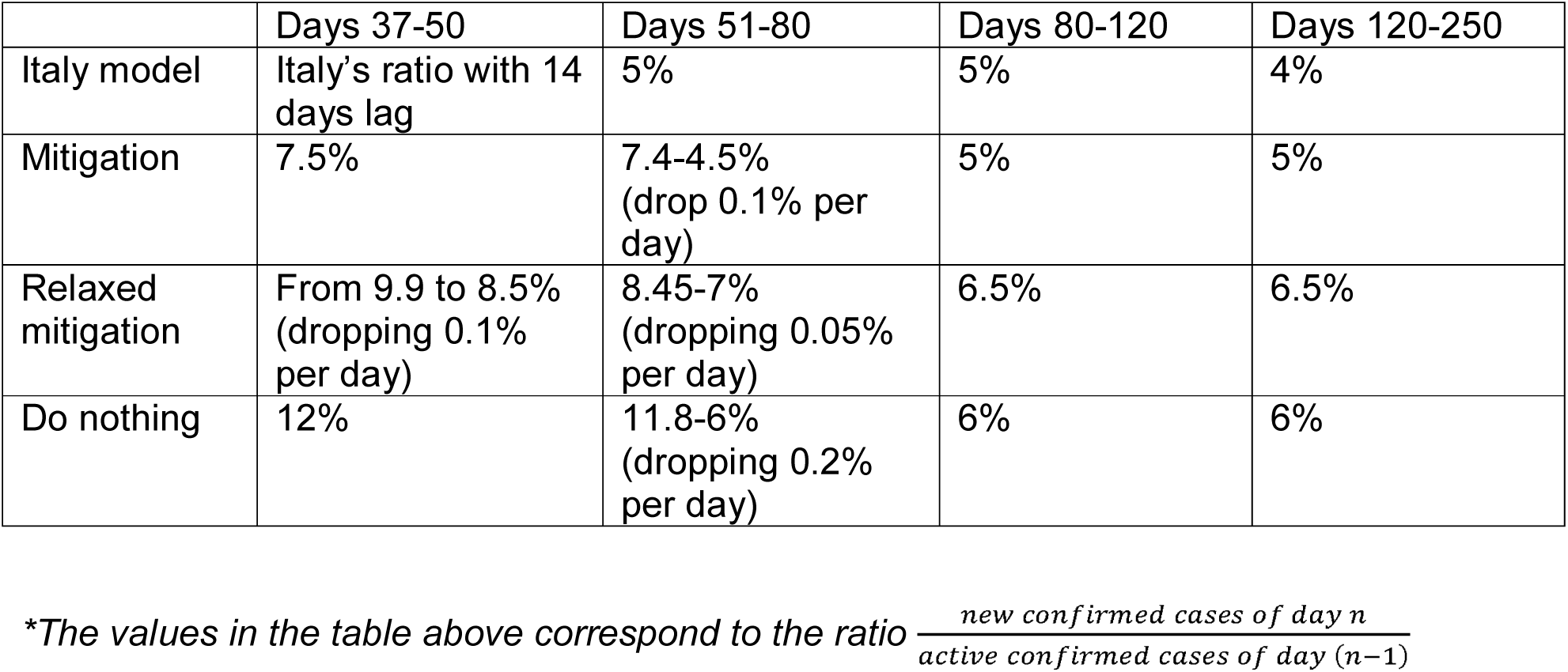
Infection growth rate at different times in the pandemic in four different scenarios*

|  | Days 37-50 | Days 51-80 | Days 80-120 | Days 120-250 |
| --- | --- | --- | --- | --- |
| Italy model | Italy's ratio with 14 days lag | 5% | 5% | 4% |
| Mitigation | 7.5% | 7.4-4.5%<br>(drop 0.1% per day) | 5% | 5% |
| Relaxed mitigation | From 9.9 to 8.5%<br>(dropping 0.1% per day) | 8.45-7%<br>(dropping 0.05% per day) | 6.5% | 6.5% |
| Do nothing | 12% | 11.8-6%<br>(dropping 0.2% per day) | 6% | 6% |

\*The values in the table above correspond to the ratio $\frac{\text{new confirmed cases of day } n}{\text{active confirmed cases of day } (n-1)}$

We assumed that an individual remains infected for 2 weeks, followed by death or immunity, and that actual cases were ∼20 times more than confirmed cases, as people with mild or no symptoms are not routinely tested(based on prior estimates of 5-to 100-fold)(18). Further details are specified in the Web appendix.

### Impact of infection rate on clinical academic workforce

We used NHS Digital data (December 2019) to quantify number of doctors in England (n=125,119)(19). Baseline number of clinical academics was estimated as 5% of doctors(20): 6255 in England. Based on UK clinical academic funding (21), we assumed 50% FTE (Full Time Equivalent) overall, equivalent to ∼3000 100% FTE academics, and 25% of doctors off sick and/or socially isolating at any time(22). There are 1953 intensive care, 7678 emergency medicine, 395 infectious diseases and 2748 respiratory doctors in England(19). We assumed doctors of any specialty could contribute to the COVID-19 academic response, and necessary research skills and training were homogeneously available throughout the medical workforce.

Clinical academics are not available for research if: (i) they are delivering frontline care due to health system strain, or (ii) they are off sick. We modelled two scenarios with no medical academic capacity at 10% (low strain on the health system) and 5% (high strain on the health system) infection rates respectively. Our outcome was the available number of medical academics during the pandemic in England. We assumed that the number of potentially available 100% FTE clinical academics in research is 3200, but it is obvious that this number has decreased from the early days of the COVID-19 pandemic. We present in detail the assumptions of modelling of available clinical academics in the Web appendix.

### Narrative analysis of case studies

1. <u>Northern Italy:</u> GQ provided first-hand experience of the pandemic as a physician in Bergamo, Italy.
2. <u>Health Care Worker (HCW) cohort study:</u> Our team recently set up and started recruitment for the “Healthcare Worker Bioresource: Immune Protection and Pathogenesis in SARS-CoV-2” Study (COVID-HCW; NCT04318314)(23).
3. <u>Nightingale Hospital:</u> A new NHS field hospital has been established, providing extra medical and intensive care capacity for provision of care to COVID-19 patients with a maximum theoretical capacity of 4000 beds, mainly intensive care(24). We describe the scenario, staff involved and clinical and research priorities, and constraints.

### Development of recommendations

Based on our model and our case studies, we have developed pragmatic recommendations for clinical research priorities relating to COVID-19.

## Results

### Population infection rate and infection growth rate

Figure 1 illustrates different scenarios of infection growth rate and population infection rate (point estimate from x-axis; and cumulative estimate from area under the curve) and the “flatten the curve” phenomenon. The higher the growth rate, the higher the peak infection rate and the quicker the system is overwhelmed by cases of COVID-19. In addition, the course of the pandemic is longer. Conversely, if the infection growth rate is reduced, the curve is “flattened” and the pandemic course is shorter. The peak infection rate will be 2.4% (14/4/20), 2.5% (28/4/20), 5.3% (23/5/20) and 16.7% (20/5/20) and the duration of this wave (from 5 March 2020) of the pandemic will be 95, 115, 240 and 240 days respectively. The cumulative infection rates correspond to the scenarios of “do-nothing” (cumulative infection rate ∼80%), “relaxed mitigation” (cumulative infection rate ∼40%), “mitigation” (cumulative infection rate ∼10%) and “Italy model” (cumulative infection rate ∼6%).

**Figure 1.**
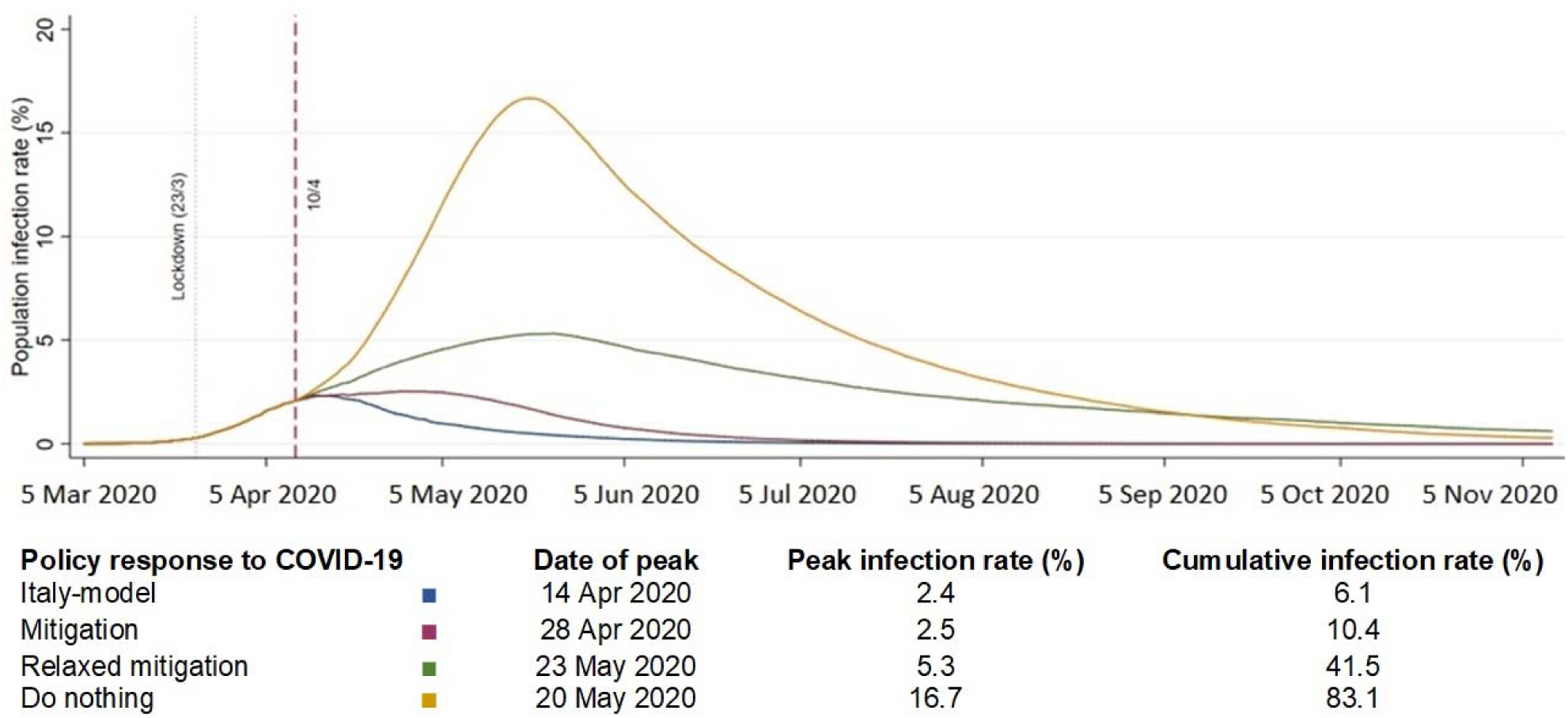
Population infection rate and daily infection growth rate in the UK during COVID-19 pandemic.

### Clinical academic capacity

Assuming the “low strain on the health system” model (where there is no academic capacity at population infection rate of 10%), Figure 2 shows that less than 400 100%FTE clinical academics (∼13%) will available after April 10 for 11, 34, 62, 76 days for the scenarios of “Italy model”, “mitigation”, “relaxed mitigation” and “do-nothing” respectively. In the “do nothing scenario”, no clinical academics are available to do research for 37 days (3/5/2020 to 8/6/2020). The predicted dates to reach 2560 clinical academics (80% normal capacity) are 23/6/2020, 1/7/2020, 3/11/2020 and 10/10/2020 for the scenarios of “Italy model”, “mitigation”, “relaxed mitigation”, and “do-nothing”, respectively.

**Figure 2:**
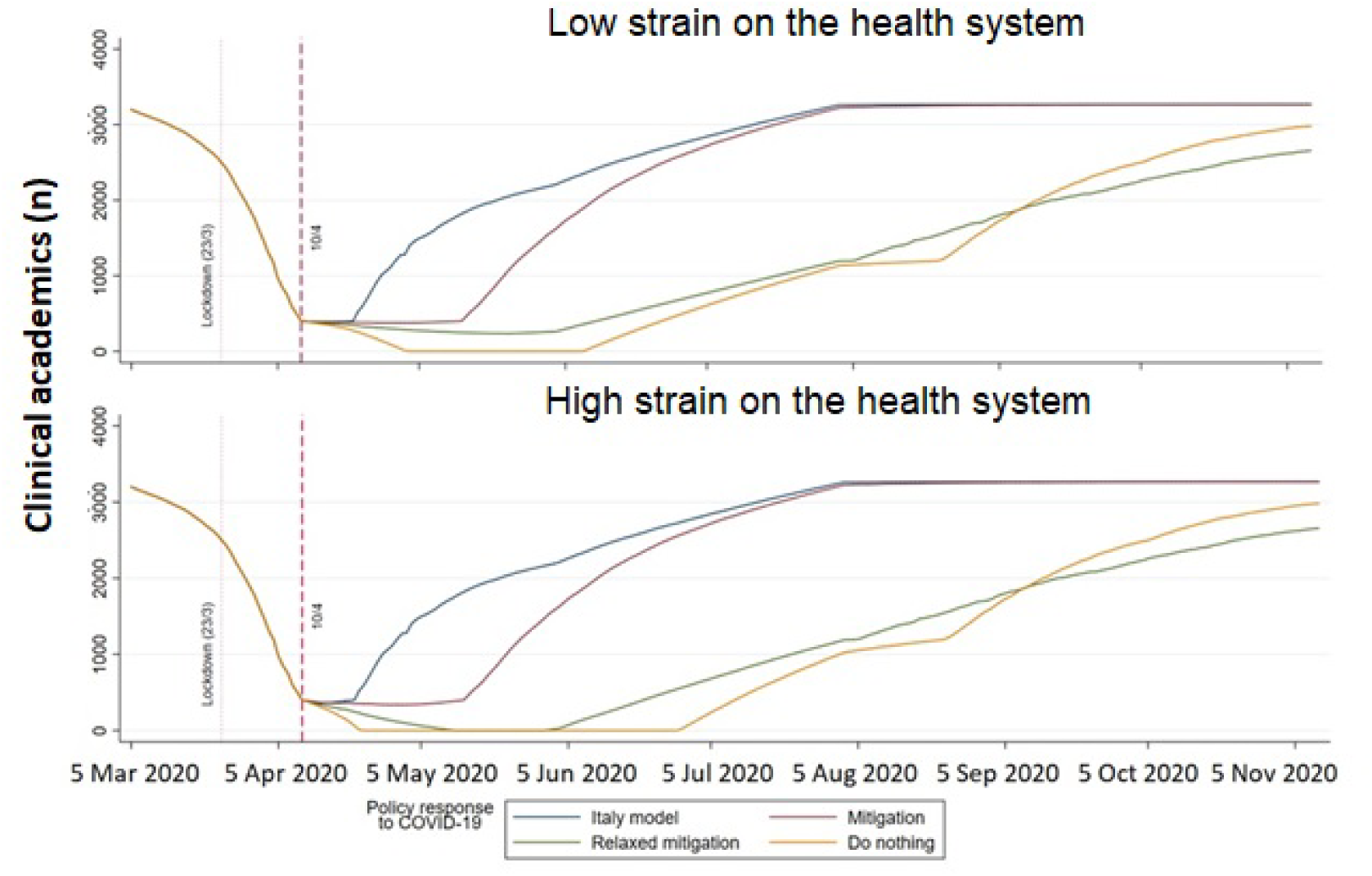
Clinical academic capacity during the COVID-19 pandemic. Low strain on health system: No clinical academic capacity at 10% population infection rate. High strain on health system: No clinical academic capacity at 5% population infection rate. Full model and assumptions in web appendix.

In the “high strain on the health system” model (where there is no academic capacity at population infection rate of 5%), in the “relaxed mitigation” scenario, no clinical academics can do research for 18 days (13/5/2020 to 30/5/2020) and in the “do-nothing” scenario, from 23/4/2020 to 28/6/2020. The predicted dates to reach 2560 clinical academics (80% normal capacity) is 23/6/2020, 2/7/2020, 7/11/2020 and 11/10/2020 for the scenarios of “Italy model”, “mitigation”, “relaxed mitigation”, and “do-nothing” respectively.

### Case studies

#### Northern Italy

*A*s the first Western region to be affected (Lombardy, Bergamo) there was effectively no warning. Almost overnight a huge surge of severely ill patients hit us.It was the beginning of a nightmare. With no treatments, we had to re-organize the hospital wards, ITU beds, transform simple general into sub-intensive units, commit all doctors from all specialities and research to COVID-19 in a matter of hours-days. It was “catastrophe medicine”, research was impossible and approaches were empiric based on analogy to other diseases. Autopsy was our only science. “Mors ubi gaudet succurrere vitae - Where the dead are happy to help the living” and we started to appreciate the high rate of thrombotic complications and pulmonary pathology, initiating empiric anticoagulation and corticosteroids. Only months later with external partnerships were formal randomized trials initiated.

#### The COVID-HCW Study

Italy informed our strategy. The team nucleus was 5 senior lecturers and 1 professor, all of whom had all their clinical work stopped as irrelevant in the pandemic (cardiac MRI) and who had a track record of monthly large grant writing and detailed systems knowledge. The hospital had no emergency department, so it was protected with a large institution behind it. “Exponential teams” were created to deliver national and local components of research permissions – permissions took 100 documents and ∼40 staff working at least part-time to deliver in 7 days (covid-consortium.com). Following scoping, we rejected all but the most basic of aspirations: to capture (questionnaire, bloods and nasal swab) 400 HCW and track changes over 16 weeks – no Clinical Trial of an Investigational Medicinal Product (CTIMP) and no direct work with COVID-19 patients. By day 16 from concept, 400 HCWs had been recruited and the study was in follow-up. At this stage, funding, aliquoting, and detailed basic science plans were embarked on (23).

#### Nightingale Hospital

The Nightingale Hospital will be the largest field hospital in Europe with the largest number of intensive care and step-down facilities for COVID-19 (Figure 3). It was set up in 14 days from initial concept to first patient admitted Nightingale is a learning system, underpinned by research. For patients, staff and wider NHS benefit, the design incorporates a commitment to learning fast and acting fast across all dimensions: clinical, operational, and staff wellbeing. Our research approach is: (i) embedded within the Quality and Learning team, (ii) simple; and (iii) high-quality and high-volume recruitment. The onsite team is backed up by QMUL, UCL and UCLP, with multidisciplinary expertise, including virology, immunology and therapeutics. From an initial two clinical academic staff (AB and JM), a research governance structure has been set up rapidly and a simple strategy has been established. COVID-19 consented studies can be observational or interventional (drugs), in patients or staff. We plan just one initial study in each domain, choosing the simplest possible approaches: patient observational(ISARIC), patient therapeutic(RECOVERY), staff observational (COVID-HCW with expansion to n=1000) and staff therapy (pre- and/or post-exposure prophylaxis studies-to be confirmed]). The first patient therapeutic trial patient will be recruited on day 8 after first patient admitted. Other studies (data, staff surveys) can be conducted at other sites (Table 1) or after the initial exponential wave peaks. In addition, there are opportunities for non-consented research, such as epidemiologic and advanced data analytics; e.g. initiatives such as DeCOVID(25) to mobilise data, computer scientists, analysts and analytic infrastructure, including and clinical expertise. There is potential to link effective learning directly to and from clinical questions.

**Figure 3:**
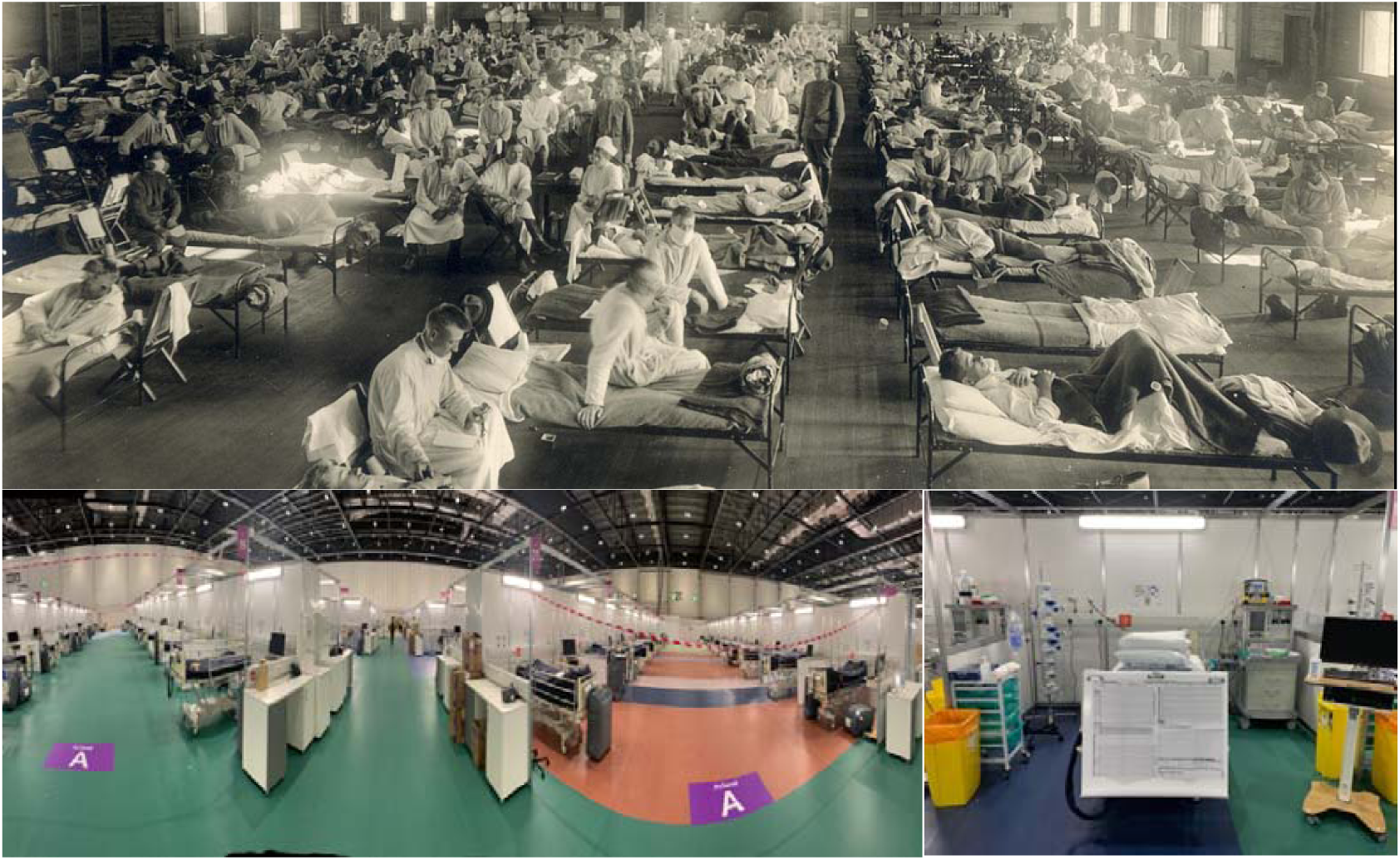
Field hospitals in pandemics: 1918 vs 2020.

**Table 1.** Simple strategy for consented studies at the Nightingale hospital, London

|  | **Patients** | **Staff** |
| --- | --- | --- |
| **Observational** | ISARIC<br>www.remapcap.org/ | HealthCare Workers study<br>www.covid-consortium.org |
| **Randomised trial** | Recovery<br>www.recoverytrial.net/<br>(later REMAP-CAP)<br>www.remapcap.org/ | (a pre-exposure prophylaxis study – 3 in preparation: we will choose one) |

### Recommendations

After discussion among co-authors, and consensus among a stakeholder group at the Nightingale Hospital, we produced recommendations for a COVID-19 clinical research strategy (Table 2).

**Table 2.**
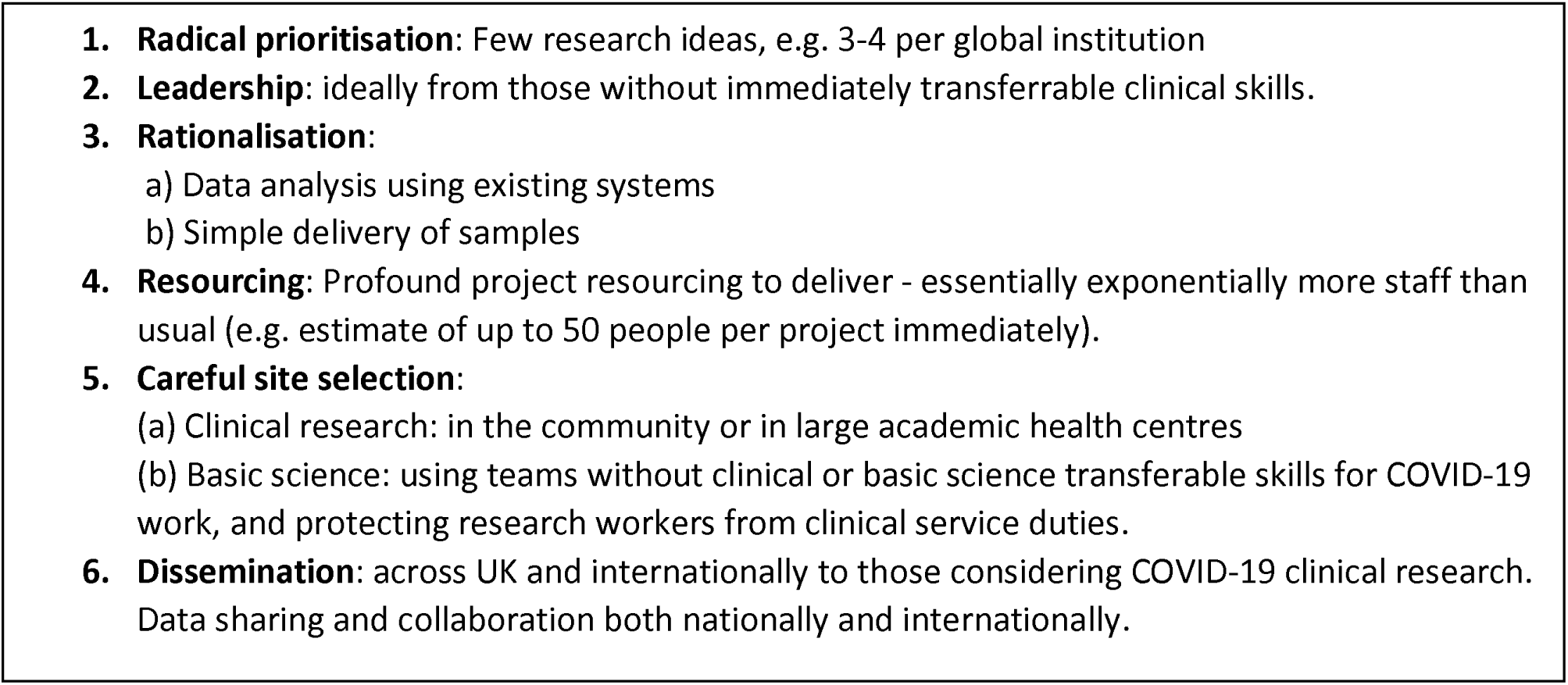
Research strategy recommendations

## Discussion

In the first study of clinical academic capacity in the COVID-19 era, we show the existential threat to research responses facing the UK and other countries. Urgent recognition and mobilisation are required to ensure prioritisation of the most appropriate and clinically imperative science. We have developed recommendations relevant to all health systems.

The healthcare and public health emergency caused by COVID-19 is not in question, fuelling global discussion, modelling and multidisciplinary research at a pace rarely seen(26-27). However, strain on clinical academic workforce and infrastructure in different countries are notable omissions. Despite programmes to promote research preparedness in epidemics, COVID-19 poses particular challenges(28) to our responses which rest ultimately on research, whether vaccines, drugs, ventilation strategies, risk prediction or machine learning. Our experiences are echoed in China, Italy and other countries facing the pandemic.

There have been quick efforts and advances in fields as diverse as genomics(29) and data science(30), with rapid-response calls from major funders(5, 31-33). However, our data signal a need for a far broader paradigm shift in research design and implementation. At every stage in the traditional research pipeline, there are roadblocks hampering swift reactions necessary to tackle COVID-19 within and across countries. Even on a “war footing” (Figure 3), research processes are unnecessarily time- and resource-consuming, particularly when involving randomized controlled studies. Specific hurdles are: (i) **Staff** - doctors and research nurses, but also access to labs; (ii) **Stuff**-consumables difficult to obtain due to challenging supply chains especially if they are competing with clinical service delivery, e.g. personal protective equipment; (iii) **Site**-ideally research space near to clinical areas; and **Systems**-approvals in a timely fashion, e.g. Research Ethics Committee, Health Research Authority, Local Research and Development team and standard operating procedures ins relevant institutions.

Emergencies as far-reaching as the current scenario require total rethinking of research delivery, and aspects that work better when some of the processes are accelerated and the permissions expedited, may well yield long-term benefits outside of COVID-19 research. Here we have modelled clinical academic time in terms of numbers of staff and time in the pandemic. However, a far deeper examination of the role of clinical academics beyond “hours at the desk” is warranted in times of public health emergency to include the “what” and “how” of their work. For example, certain tasks such as research permissions and data analysis may be diverted away from clinical academics, who may be better placed to act as conduits between the clinical and public health spheres and teams of non-clinical researchers. The needs of the hour are patient-centred, data-driven and time-responsive, and it may be time to usefully change the role and function of the clinical academic. It is worth noting that this is occurring against a backdrop of declining clinical academic numbers(21).

Our simulations suggest the pandemic will create health system strain for many critical months. Depending on a range of COVID-related factors, we show that the clinical academic workforce may be depleted when it is needed most to lead and conduct clinical research, even in a relatively well-resourced context such as the UK, whether by funding, number of universities and staff, infrastructure or policy. Therefore, other countries are likely to be worse affected. COVID-19 research is least likely to occur where it is most needed, magnifying the well-documented “10-90 research gap”, where only 10% of resources for global healthcare research are devoted to low-income settings where 90% of preventable deaths occur(34). Although COVID-19 is a unique threat, there are lessons to be learned from prior health research strategies to address structural inequities, such as the Global Fund for Malaria, TB and HIV/AIDS(29). Without coordinated international responses, including urgent funding and infrastructure, research will be retrospective, patchy and unlikely to have an effect.

We provide six clear recommendations for science in the UK and globally in relation to COVID-19 (Radical prioritisation, Leadership, Rationalisation, Resourcing, Careful site selection and Dissemination). Radical prioritisation is important where field hospitals are being established in rapid timescales in different countries with delivery constraints. High-quality evidence can be obtained, but studies need to be lean with minimal complexity for key operational steps: consenting, randomisation, drug delivery, monitoring, outcomes and follow-up. The number of patients recruited to deliver definitive answers needs to be large, with fast recruitment across multiple sites. Furthermore, adaptive trial designs are preferred as new arms (e.g. multi-drug) can be generated swiftly and other arms dropped (e.g. supportive care if one arm has a signal of efficacy) without restarting permissions, via substantial amendments(38, 39).

Leadership and rationalisation are the next key steps. Balance needs to be struck between clinical researchers in contact with the “frontline” so that research questions are clinically relevant and timely, and having research active leaders who will not be protected from frontline work. Rationalisation involves a study selection strategy that is deeply resourced for a limited number (1 or 2) studies per COVID cohort. In selecting these studies, a single study of one investigational medical product versus standard of care (supportive care) with 50:50 randomisation is inefficient compared to studies with multiple therapeutic arms. Most single agent approaches to COVID-19 are likely to have, at most, a modest effect.

We used a stochastic model accounting for infection rate, infection growth rate and clinical academic capacity using up-to-date official statistics. There are limitations to our model and its assumptions. Our model was simple and was based only on observational patterns of the number of new cases and actual cases from publicly available data(17). We conducted analyses on 10 April, and on 11 April, some extra ∼3000 cases were added retrospectively and distributed over the past 10 days-we did not include these data. It did not take into account infectious disease epidemiology parameters, such as the basic reproductive number (R0), and we did not consider differing levels of risk of infection(35-37). Our model on the availability of clinical academics makes several assumptions (Web Appendix), including the total number of 100%FTE academics as ∼3200, with a uniform skillset across the workforce.

## Conclusions

In the first study to model and estimate the impact of COVID-19 on the coordinated clinical academic response at system level, we show that all countries face depletion of their clinical academic workforce for several months, which will greatly hamper research in prevention and treatment. The number of studies needs to be rationalised urgently and background problems in clinical academia need to be overcome quickly. To quote Sir Jeremy Farrar, “The only exit from this pandemic is through science”(40) and that requires staffing.

## Contributorship

Research question: JCM and AB.

Study design and analysis plan: AB, JCM, MK.

Statistical analysis: MK, AB

Drafting initial and final versions of manuscript: AB, JM

Critical review of early and final versions of manuscript: all authors, specifically: clinical (AB, JCM, CM, TT, GQ, JC), infectious disease (MN), data science (AGL), informatics (SD), and public health(HH).

## Data Availability

There are no data available in relation to this manuscript.

## Web appendix

### Simulation study for population infection rate

Our simulation with 640 000 hypothetical individuals (1/100 of the population in the UK) started on the 5 of March, when 23 people had COVID-19 and the rest were healthy(17). We selected 23, because we selected the first date that the confirmed cases were >100 in worldometer (116 cases on the 5/3). So, we assume that on that date, we had 116*20∼= 2300 cases, so we used in our simulation 2300/100=23.

The probability *P*(new case at day n) is equal to

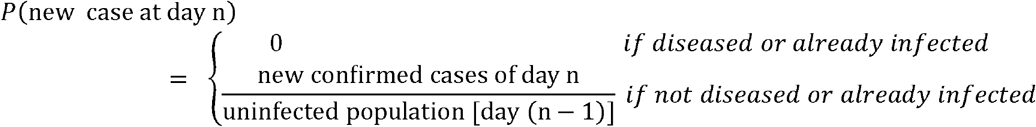

However, we have that 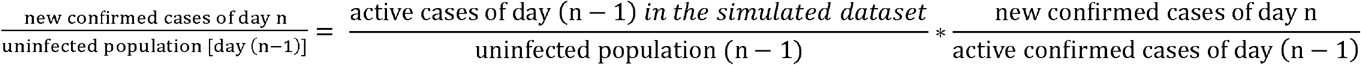

We inserted values for the ratio 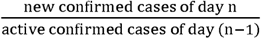 from official statistics(17) from day 1 (March 5) to day 36 (April 10). Then we assumed different scenarios that are illustrated in web table 1.

### Effect of the population infection rate on the medical academic capacity

We made the following assumptions:

1. The number of academics dropped linearly from 3200 to 2400 between day 0 and day 36, because 800 (25% were self-isolated on the 10 April). We further assumed that 800 academics will be quarantined from day 36 to day 90 and will return to work linearly until day 150.
2. The number of clinical academics available on 10 April was 400.
3. We modelled two scenarios for medical academic capacity at 10% (low strain on health system) and 5% (high strain on health system) infection rates respectively, assuming that 1000 clinical researchers would have to work in hospital per 1% of active cases in the population, if the active cases are <2.07%, i.e. infection rate of the 10^th^ of April). In the 10% and 5% cut-off models respectively, we assumed ∼50 (i.e. 400/7.93) 2) and ∼136 (i.e. 400/2.93) extra researchers can be available per 1% of active cases (≥2.07% i.e. infection rate on 10 April).

## References

1. Hick JL, Biddinger PD. Novel Coronavirus and Old Lessons - Preparing the Health System for the Pandemic. N Engl J Med. 2020 Mar 25. doi: 10.1056/NEJMp2005118. [Epub ahead of print]

2. Li Q, Guan X, Wu P, Wang X, Zhou L, Tong Y, Ren R, Leung KSM, Lau EHY, Wong JY, Xing X, Xiang N, Wu Y, Li C, Chen Q, Li D, Liu T, Zhao J, Liu M, Tu W, Chen C, Jin L, Yang R, Wang Q, Zhou S, Wang R, Liu H, Luo Y, Liu Y, Shao G, Li H, Tao Z, Yang Y, Deng Z, Liu B, Ma Z, Zhang Y, Shi G, Lam TTY, Wu JT, Gao GF, Cowling BJ, Yang B, Leung GM, Feng Z. Early Transmission Dynamics in Wuhan, China, of Novel Coronavirus-Infected Pneumonia. N Engl J Med. 2020 Mar 26;382(13):1199–1207.

3. Zhou F, Yu T, Du R, Fan G, Liu Y, Liu Z, Xiang J, Wang Y, Song B, Gu X, Guan L, Wei Y, Li H, Wu X, Xu J, Tu S, Zhang Y, Chen H, Cao B. Clinical course and risk factors for mortality of adult inpatients with COVID-19 in Wuhan, China: a retrospective cohort study. Lancet. 2020 Mar 28;395(10229):1054–1062. doi: 10.1016/S0140-6736(20)30566-3. Epub 2020 Mar 11.

4. Kandel N, Chungong S, Omaar A, Xing J. Health security capacities in the context of COVID-19 outbreak: an analysis of International Health Regulations annual report data from 182 countries. Lancet. 2020 Mar 28;395(10229):1047–1053. doi: 10.1016/S0140-6736(20)30553-5. Epub 2020 Mar 18.

5. Wellcome Trust. Wellcome statements on novel coronavirus (COVID-19). https://wellcome.ac.uk/press-release/wellcome-statements-novel-coronavirus-covid-19 (Accessed 5 April 2020)

6. Song P, Karako T. COVID-19: Real-time dissemination of scientific information to fight a public health emergency of international concern. Biosci Trends. 2020 Mar 16;14(1):1–2. doi: 10.5582/bst.2020.01056. Epub 2020 Feb 25.

7. Haffajee RL, Mello MM. Thinking Globally, Acting Locally - The U.S. Response to Covid-19. N Engl J Med. 2020 Apr 2. doi: 10.1056/NEJMp2006740. [Epub ahead of print]

8. Kuhlmann E, Batenburg R, Wismar M, Dussault G, Maier CB, Glinos IA, Azzopardi-Muscat N, Bond C, Burau V, Correia T, Groenewegen PP, Hansen J, Hunter DJ, Khan U, Kluge HH, Kroezen M, Leone C, Santric-Milicevic M, Sermeus W, Ungureanu M.A call for action to establish a research agenda for building a future health workforce in Europe. Health Res Policy Syst. 2018 Jun 20;16(1):52. doi: 10.1186/s12961-018-0333-x.

9. Rees MR, Bracewell M; Medical Academic Staff Committee of the British Medical Association. Academic factors in medical recruitment: evidence to support improvements in medical recruitment and retention by improving the academic content in medical posts. Postgrad Med J. 2019 Jun;95(1124):323–327. doi: 10.1136/postgradmedj-2019-136501. Epub 2019 Jun 8.

10. Royal College of Physicians. COVID-19 and its impact on NHS workforce. 5 April 2020. https://www.rcplondon.ac.uk/news/covid-19-and-its-impact-nhs-workforce (Accessed 5 April 2020)

11. University College London. Clinical academics freed up to support NHS during Covid-19 pandemic. 18 March 2020. https://www.ucl.ac.uk/news/2020/mar/clinical-academics-freed-support-nhs-during-covid-19-pandemic (Accessed 5 April 2020)

12. Zhang Z, Liu S, Xiang M, Li S, Zhao D, Huang C, Chen S. Protecting healthcare personnel from 2019-nCoV infection risks: lessons and suggestions. Front Med. 2020 Mar 23. doi: 10.1007/s11684-020-0765-x. [Epub ahead of print]

13. The Lancet. COVID-19: protecting health-care workers. Lancet. 2020 Mar 21;395(10228):922. doi: 10.1016/S0140-6736(20)30644-9.

14. Banerjee A, Pasea L, Harris S, Gonzalez-Izquierdo A, Torralbo A, Shallcross L, Noursadeghi M, Pillay D, Sebire N, Holmes C, Pagel C, Wong WK, Langenberg C, Williams B, Denaxas S, Hemingway H. Estimating excess 1-year mortality from COVID-19 according to underlying conditions and age in England: a population based cohort using NHS health records in 3.8 million adults. 2020. Under review.

15. Denaxas S, Gonzalez-Izquierdo A, Direk K, Fitzpatrick NK, Fatemifar G, Banerjee A, Dobson RJB, Howe LJ, Kuan V, Lumbers RT, Pasea L, Patel RS, Shah AD, Hingorani AD, Sudlow C, Hemingway H. UK phenomics platform for developing and validating electronic health record phenotypes: CALIBER. J Am Med Inform Assoc. 2019;26(12):1545–59.

16. Kuan V, Denaxas S, Gonzalez-Izquierdo A, Direk K, Bhatti O, Husain S, Sutaria S, Hingorani M, Nitsch D, Parisinos CA, Lumbers RT, Mathur R, Sofat R, Casas JP, Wong ICK, Hemingway H, Hingorani AD.A chronological map of 308 physical and mental health conditions from 4 million individuals in the English National Health Service. Lancet Digit Health. 2019 May 20;1(2):e63–e77. doi: 10.1016/S2589-7500(19)30012-3.

17. Worldometer. COVID-19 coronavirus pandemic country estimates: UK. 5 April 2020. https://www.worldometers.info/coronavirus/country/uk/ (Accessed 5 April 2020)

18. Ioannidis, JPA. A fiasco in the making? As the coronavirus pandemic takes hold, we are making decisions without reliable data. StatNews. March 17 2020. https://www.statnews.com/2020/03/17/a-fiasco-in-the-making-as-the-coronavirus-pandemic-takes-hold-we-are-making-decisions-without-reliable-data/

19. NHS Digital Workforce statistics. December 2019. https://digital.nhs.uk/data-and-information/publications/statistical/nhs-workforce-statistics/december-2019 (Accessed 5 April 2020)

20. Health Education England. Health careers. Clinical academic medicine. https://www.healthcareers.nhs.uk/explore-roles/doctors/career-opportunities-doctors/clinical-academic-medicine (Accessed 5 April 2020)

21. Medical Schools Council. Survey of Medical Clinical Academic Staffing Levels 2018. https://www.medschools.ac.uk/media/2491/msc-clinical-academic-survey-report-2018.pdf

22. Hope R. Coronavirus: One in four NHS doctors ‘sick or in isolation’. https://news.sky.com/story/coronavirus-one-in-four-nhs-doctors-sick-or-in-isolation-11965886

23. COVID-19: Healthcare Worker Bioresource: Immune Protection and Pathogenesis in SARS-CoV-2 (COVID19-HCW) https://clinicaltrials.gov/ct2/show/NCT04318314

24. BBC News. Coronavirus: Nightingale Hospital opens at London’s ExCel centre. 3 April 2020. https://www.bbc.co.uk/news/uk-52150598.

25. Alan Turing Institute. Call for COVID-19 rapid response data science taskforce. https://www.turing.ac.uk/work-turing/research-and-funding-calls/call-covid-19-rapid-response-data-science-taskforce

26. Weissman GE, Crane-Droesch A, Chivers C, Luong T, Hanish A, Levy MZ, Lubken J, Becker M, Draugelis ME, Anesi GL, Brennan PJ, Christie JD, Hanson Iii CW, Mikkelsen ME, Halpern SD. Locally Informed Simulation to Predict Hospital Capacity Needs During the COVID-19 Pandemic. Ann Intern Med. 2020 Apr 7. doi: 10.7326/M20-1260.

27. Shoukat A, Wells CR, Langley JM, Singer BH, Galvani AP. Projecting demand for critical care beds during COVID-19 outbreaks in Canada. CMAJ. 2020 Apr 8. pii: cmaj.200457.

28. International Severe Acute Respiratory and emerging Infection Consortium. https://isaric.tghn.org/

29. Shen Z, Xiao Y, Kang L, Ma W, Shi L, Zhang L, Zhou Z, Yang J, Zhong J, Yang D, Guo L, Zhang G, Li H, Xu Y, Chen M, Gao Z, Wang J, Ren L, Li M. Genomic diversity of SARS-CoV-2 in Coronavirus Disease 2019 patients. Clin Infect Dis. 2020 Mar 4. pii: ciaa203. doi: 10.1093/cid/ciaa203

30. Yang Z, Zeng Z, Wang K, Wong SS, Liang W, Zanin M, Liu P, Cao X, Gao Z, Mai Z, Liang J, Liu X, Li S, Li Y, Ye F, Guan W, Yang Y, Li F, Luo S, Xie Y, Liu B, Wang Z, Zhang S, Wang Y, Zhong N, He J. Modified SEIR and AI prediction of the epidemics trend of COVID-19 in China under public health interventions. J Thorac Dis. 2020 Mar;12(3):165–174.

31. Kiefer S, Knoblauch AM, Steinmann P, Barth-Jaeggi T, Vahedi M, Maher D, Utzinger J, Wyss K. Operational and implementation research within Global Fund to Fight AIDS, Tuberculosis and Malaria grants: a situation analysis in six countries. Global Health. 2017 Mar 24;13(1):22. doi: 10.1186/s12992-017-0245-5.

32. Gates Foundation. Announcing the COVID-19 Therapeutics Accelerator. https://www.gatesfoundation.org/TheOptimist/Articles/coronavirus-mark-suzman-therapeutics

33. National Institute of Health. Coronavirus news, funding and resources for global health researchers.

34. Lown B, Banerjee A. The developing world in The New England Journal of Medicine. Global Health. 2006 Mar 16;2:3.

35. Prem K, Liu Y, Russell TW, Kucharski AJ, Eggo RM, Davies N; Centre for the Mathematical Modelling of Infectious Diseases COVID-19 Working Group, Jit M, Klepac P. The effect of control strategies to reduce social mixing on outcomes of the COVID-19 epidemic in Wuhan, China: a modelling study. Lancet Public Health. 2020 Mar 25. pii: S2468-2667(20)30073-6

36. Kucharski AJ, Russell TW, Diamond C, Liu Y, Edmunds J, Funk S, Eggo RM; Centre for Mathematical Modelling of Infectious Diseases COVID-19 working group. Early dynamics of transmission and control of COVID-19: a mathematical modelling study. Lancet Infect Dis. 2020 Mar 11. pii: S1473-3099(20)30144-4.

37. Verity R, Okell LC, Dorigatti I, Winskill P, Whittaker C, Imai N, Cuomo-Dannenburg G, Thompson H, Walker PGT, Fu H, Dighe A, Griffin JT, Baguelin M, Bhatia S, Boonyasiri A, Cori A, Cucunubá Z, FitzJohn R, Gaythorpe K, Green W, Hamlet A, Hinsley W, Laydon D, Nedjati-Gilani G, Riley S, van Elsland S, Volz E, Wang H, Wang Y, Xi X, Donnelly CA, Ghani AC, Ferguson NM. Estimates of the severity of coronavirus disease 2019: a model-based analysis. Lancet Infect Dis. 2020 Mar 30. pii: S1473-3099(20)30243-7.

38. Randomised Evaluation of COVID-19 thERapY (RECOVERY trial). https://www.recoverytrial.net/ http://www.isrctn.com/ISRCTN50189673

39. A Randomised, Embedded, Multi-factorial, Adaptive Platform Trial for Community-Acquired Pneumonia (REMAP-CAP trial) https://www.remapcap.org/

40. Farrar J. Opinion: The only exit from this pandemic is through science. We must fund it. Globe and Mail. 6 April 2020. https://www.theglobeandmail.com/opinion/article-the-only-exit-from-this-pandemic-is-through-science-we-must-fund-it/

